# Pattern and risk factors of menstrual regulation service use among ever-married women in Bangladesh: evidence from a nationally representative cross-sectional survey

**DOI:** 10.1101/2022.03.09.22272531

**Authors:** Md. Rashed Alam, Md. Nuruzzaman Khan, Yothin Sawangdee

**Author notes:** Corresponding author Yothin Sawangdee, ^3^Institute for Population and Social Research, Mahidol University, Thailand.

## Abstract

**Background:** Around 47% of the total conceptions in Bangladesh are unintended which leads to several adverse consequences, including maternal and child mortality. Availability of menstrual regulation (MR) service and its use can help women to end conception at an earlier stage, as such, reducing adverse consequences related to the unintended pregnancy. We explored the prevalence and determinants of MR service knowledge and its use among ever-married women in Bangladesh.

**Methods:** A total of 20 127 ever-married women data from the 2017 Bangladesh Demographic and Health Survey were analyzed. Knowledge about menstrual regulation (MR) and its use were our outcomes of interest. Several individual, household and community-level factors were considered as explanatory variables. The multilevel mixed-effects Poisson regression model was used to determine the factors associated with MR service knowledge and its use in Bangladesh.

**Results:** Around 71% of the total analyzed women reported they know about MR service while only 7% of them reported they used this service within three years of the survey date. MR service knowledge was found to be higher among women with increased age and education and engaged in income-generating employment. Knowledge about MR service was also found to be higher among women whose husbands were higher educated and engaged in physical work or business. Rural women and women who resided in the community with lower poverty and higher illiteracy were reported lower knowledge of MR service. MR service use was found higher among higher-aged women, women whose husbands were either physical workers or businessmen, women who have an increased number of children and inherent in the community with lower poverty. Lower use of MR service was found among women who resided in the Chattogram, Khulna, and Mymensingh divisions and women who resided in the community with increased illiteracy.

**Conclusion:** Use of MR service is very low in Bangladesh. This could be responsible for higher adverse consequences related to unintended pregnancy including higher maternal and child mortality. Policies and programs are important to aware women of MR.

## Introduction

Bangladesh, the 8^th^ largest populated country in the world, has around 20 million unintended conceptions every year that equates to around 47% of the country’s total conceptions. Around half of them are ended before live births, mostly through unsafe abortion outside the formal healthcare facilities, as abortion is legally prohibited in Bangladesh unless it is required to save a woman’s life. The remaining unintended conception are ended through life births, which contribute to around 25% of the total live births in the year in Bangladesh. Both unsafe abortion and live birth as a result of unintended conception are found to be associated with adverse birth and maternal health outcomes in low- and middle-income countries (LMICs), including Bangladesh (UNICEF 2019 & UN 2021). They include but not limited to pregnancy complications, stillbirths, preterm birth, low birth weight, neonatal, under-five, and maternal mortality. These are challenges to achieving the Sustainable Development Goals (SDGs) 3; health and wellbeing for all; particularly, to achieve the SDGs’ targets to reduce neonatal (12 per 1,000), under-five (25 per 1,000) and maternal (73 per 10,0000 live births) mortality by 2030 (UN-SDGs Goals 3).

Menstrual regulation (MR), a type of abortion that is performed in a very early stage of pregnancy, has been used in Bangladesh as an alternative of abortion to end pregnancy due to unintended conceptions. The government of Bangladesh in 1979 was approved MR as part of the family planning services and now available in governmental healthcare facilities. It combines manual vacuum aspiration and mifepristone/ misoprostol to regulate the menstrual cycle within 10/12 weeks of conception (Benson et al., 2011; Hossain et al., 2017 & Guttmacher Institute 2017).

Though MR service is available in the governmental healthcare facilities in free of cost or lower cost to access, however, such service use is still very low. The use of MR service is even now declining, alternative to abortion, which use is now increasing (Huda et al. 2010; S. Singh et al., 2017; Guttmarch Institute, 2012). Consequently, abortion related adverse consequences, including maternal mortality, are very higher, which create a substantial burden to the healthcare system. For instance, abortion related maternal mortality rate was increased 35.5 per 1,000 live births in 2016 (7% of the total maternal mortality) 18 per 1,000 live births in 2010 (1% of the total maternal mortality) (Hossain A et al., 2014; M. Singh et al., 2019; Rana et al., 2019). Further policies and programs are therefore important to increase access of MR services. For this, proper understanding of the current level of knowledge of MR and the factors associated with MR service use are important. However, this understanding is still lacking. So far, only one research is published by using the nationally representative 2014 BDHS data which explored the basic socio-demographic factors associated with the MR service use. This is line of the other studies conducted in LMICs (Holmes K et al, 2021; Chae et al., 2017). However, as far know no research has been conducted yet in Bangladesh to current level of knowledge on MR service use. In addition, age-standardized prevalence knowledge of MR service and MR service use have not yet been investigated.

Therefore, the aims of this study were three-fold: (i) to determine the age-adjusted prevalence of MR service use and knowledge about MR, (ii) to explore the factors associated with MR service knowledge and (iii) to explore the factors associated with MR service use.

## Methods

### Study design and sample

The data analysed were extracted from the 2017 Bangladesh Demographic Health Survey (BDHS 2017). It is a nationally representative survey conducted in every three years by following a two-stage stratified random sampling method. The National Institute of Population Research and Training (NIPORT) conducted this survey. Developments partners, including the Macro International, ICF international, Mitra and Associates, US Agency for International Development (USAID), and Ministry of Health and Family Welfare, Bangladesh provided technical and financial supervision. In the first stage of sampling, 675 PSUs were randomly selected from 293 579 PSUs which were generated by the Bangladesh Bureau of Statistics as part of conducting the 2011 National Population Census (BBS, 2014; BDHS, 2017). Data were collected from the 672 PSUs of them, whereas 20 160 households were selected for data collection with 30 households from each selected PSU in the second stage of sampling. Of them data collection was undertaken in 19 457 households with a 96.5% inclusion rate. There were 20 127 ever married women interviewed in from these selected households. Of them, we analysed 14 346 women data knowledge about MR and 1056 women use MR service in the 3 years preceding the survey (Figure. 1).

**Figure 1:**
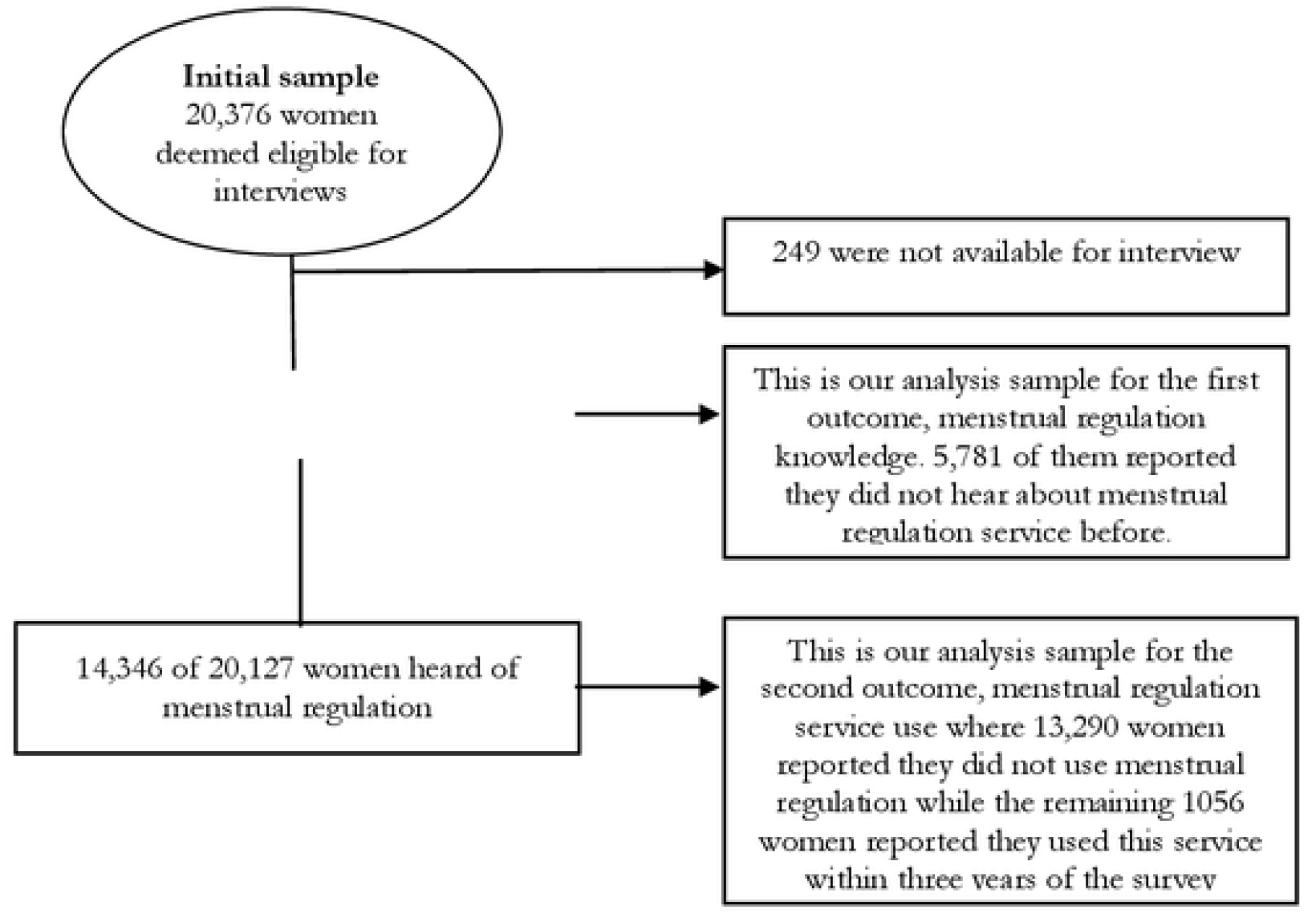
Study sample selection pcocess: Bangladesh Demographic and Health Survey 2017-18

### Outcome variable

Respondents’ knowledge on MR and MR service use status within three years of the survey date were our outcomes of interest. Knowledge related data were collected by asking women *“have you ever heard of menstrual regulation (MR)?”*. The woman who answered positively to this item was then asked *“have you ever used MR?”*. Responses were recorded dichotomously for both questions (yes, no).

### Predictor variables

The variables that were found significant for MR knowledge and MR service use were considered as predictor variables. For this, we first conducted an extensive literature search in five databases (PubMed, Embase, CINHAL, Web of Science and Goggle Scholar) and significant variables were listed. Their availability in the dataset we analysed were then checked and significance were tested. Finally, significant variables were kept in our analysis classified under three broad heading following the socio-ecological model of health: individual level factors, household level factors, and community level factors (Bronfenbrenner, U. 1994; Dahlberg & Krug, 2006). Individual level factors were women’s age (15-19 years, 20-34 years and ≥35 years), education (no education, primary education, secondary education and higher education), age at marriage (below 18 years and 18 years and above) and working status (yes, no). Household level factors were partners’ education (no education, primary education, secondary education and higher education), occupation (agricultural, physical workers, services, business and others), number of children ever born (no child, 1-2 children and >2 children) and wealth quintile (poorest, poorer, middle, richer and richest). Place of residence (urban and rural), region of residence (Barisal. Chattogram, Dhaka, Khulna, Mymensigh, Rajshahi, Rangpur and Sylhet), community level poverty (high poverty, moderate poverty, low poverty and middle to richest community) and community level illiteracy (low, moderate and high) were considered as community level factors.

### Statistical Analysis

Descriptive statistics were used to describe the characteristics of the respondents. Age-standardized prevalence of MR service knowledge and its use with their 95% confidence interval (95% CI) were also calculated. We calculated age-standardised estimates to the 2011 Census population of Bangladesh using the direct method [∑*r*_*i*_ X *P*_*i*_/∑*P*_*i*_], where *r*_*i*_ is the prevalence of MR service knowledge or use in age group *i* and *Pi* is the population size in the *i*th age group. Predictors of MR service knowledge and its use status were determined using the multilevel mixed effect Poisson regression model. The reason of using this model were nested structure of the data we analysed and higher prevalence of MR use where conventional logistics regression model produces incorrect results ((Al-Balushi et al., 2020; Nelder, J. A, 1989). Five models were run separately for each outcome by following the progressive model building technique. Model 1 was null model determined the overall variability of MR service knowledge and its use across the PSUs. Individual level factors were adjusted with model 2. Household level factors were adjusted in model 3. Community level factors were considered in model 4. Model 5 was the final model where all individual, household and community level factors were adjusted. Sampling weight was considered in all analyses. Results were reported as Prevalence Ratio (PR) with its 95% CI. All analyses were performed using statistical software packages Stata (version 13·0; Stata Corp LP, College Station, Texas).

### Ethics statement

The study analyzed secondary data of BDHS. The BDHS is a part of the worldwide Demographic and Health Surveys (DHS) programme. The Bangladesh Ministry of Health and Family Welfare provided ethical approval for the surveys. The National Institute of Population Research and Training conducted the surveys with the support of the United States Agency for International Development (USAID). The survey protocol was approved by the National Research Ethics Committee in Bangladesh and ORC Macro (Macro International Inc.) Institutional Review Board. Informed consent was obtained from all participants.

### Results

Of the 20 127 women data that were analysed in this study, 14 346 women reported they know about MR service which equates to 71% of the total respondents (Table 1). Of the women who reported positively, only 7% use MR service within three years of the survey date. The mean age of the respondents was 31.59 years at the time of the survey and mean years of education was 5.77 years.

**Table 1:** Background characteristics of the respondents

| **Variables** | **Mean (SE) or Frequency (%)** |
| --- | --- |
| Women's age at the time of survey (mean±SD) | 31.59 (±9.18) |
| Women's age at marriage | 16.15 (±2.97) |
| Woman's education | 5.77±4.14 |
| Heard about menstrual regulation |  |
| Yes | 14346 (71.28) |
| No | 5781 (28.72) |
| Use menstrual regulation within three years of the survey |  |
| Yes | 1056 (7.36) |
| No | 13290 (92.64) |

Of the women who used MR services, 62% resided in rural area (Figure 2). Moreover, of the urban women who know about MR service, only 9% used and about 93% of the rural women did not use of MR service within three years of the survey date. In contrary, nearly 7% of the total rural women who know about MR services used, MR services within three years of the survey date.

**Figure 2:**
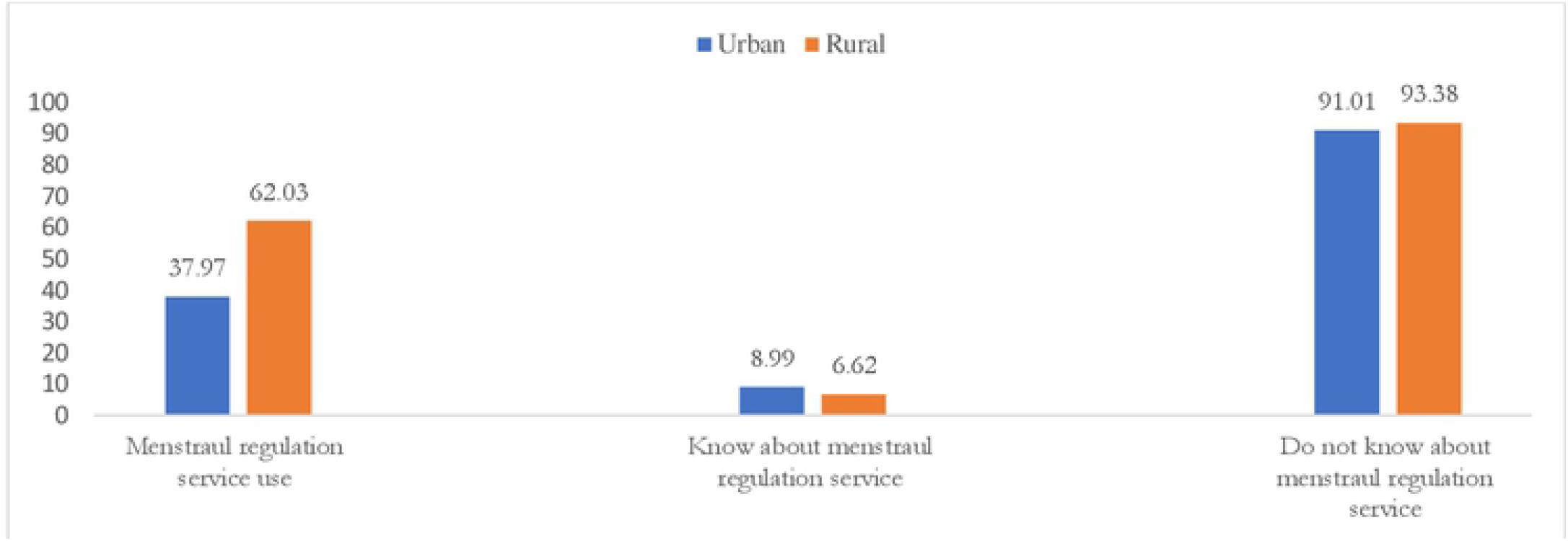
Menstrual regulation use status and knowledge by place of residence of BDHS 2017.

### Prevalence of knowledge about Menstrual Regulation and MR service use

We calculated age-adjusted prevalence of MR service knowledge and use of MR service across individual-, household, and community level factors (Table 2). MR service knowledge and MR service use were found to be higher among respondents with increased age, education and who got marriage after their ages of 18 years or later. Women with formal engagement of employment were reported more use and knowledge of MR service. We found increase knowledge of MR and MR service use among women whose partners were educated and engaged in either services or business. Urban women reported more knowledge and use of MR. Both knowledge and use of MR were found higher among women who resided in the community with higher literacy and lower poverty.

**Table 2:** Age-adjusted prevalence of knowledge and use of menstrual regulation service across individual-, household-, and community level characteristics, Bangladesh.

| Characteristics | Knowledge about menstrual regulation service % (95% CI) | Menstrual regulation service use % (95% CI) |
| --- | --- | --- |
| **Age of respondents** |  |  |
| 15-19 | 55.34 (52.24 – 58.40) | 1.56 (0.95 – 2.55) |
| 20-34 | 74.10 (72.30 – 75.82) | 6.53 (5.89 – 7.22) |
| ≥35 | 71.67 (69.53 – 73.61) | 9.82 (8.90 – 10.83) |
| **Respondent educational level** |  |  |
| No education | 61.46 (58.61 – 64.23) | 6.90 (5.68 – 8.37) |
| Primary education | 68.66 (66.46 – 70.78) | 8.13 (7.21 – 9.14) |
| Secondary education | 73.16 (71.25 – 74.98) | 6.90 (6.24 – 7.63) |
| Higher education | 84.80 (82.90 – 86.52) | 7.49 (6.25 – 8.95) |
| **Age at first marriage** |  |  |
| Early marriage (below 18 years) | 69.75 (67.87 – 71.57) | 7.51 (6.92 – 8.15) |
| Marriage 18 years and above | 75.93 (73.88 – 77.87) | 6.93 (5.99 – 7.99) |
| **Respondent currently working status** |  |  |
| No | 68.66 (66.47 – 70.81) | 7.15 (6.42 – 7.95) |
| Yes | 74.13 (72.31 – 75.86) | 7.57 (6.86 – 8.35) |
| **Husband educational status** |  |  |
| No education | 64.96 (62.16 – 67.66) | 6.50 (5.50 – 7.67) |
| Primary education | 68.69 (66.48 – 70.82) | 7.13 (6.25 – 8.12) |
| Secondary education | 73.77 (71.78 – 75.66) | 8.11 (7.17 – 9.15) |
| Higher education | 82.96 (81.12 – 84.66) | 7.95 (6.71 – 9.39) |
| **Husband occupational status** |  |  |
| Agriculture | 67.63 (65.08 – 70.07) | 6.26 (5.41 – 7.23) |
| Physical workers | 70.84 (68.82 – 72.77) | 7.14 (6.37 – 7.99) |
| Service | 84.84 (81.92 – 87.36) | 8.69 (6.75 – 11.13) |
| Business | 75.74 (73.34 – 77.99) | 9.22 (8.10 – 10.46) |
| Others | 63.74 (45.66 – 78.62) | 6.32 (1.35 – 24.87) |
| **Wealth Index** |  |  |
| Poorest | 64.09 (61.04 – 67.03) | 4.88 (3.97 – 5.99) |
| Poorer | 68.41 (65.78 – 70.92) | 6.71 (5.61 – 8.00) |
| Middle | 70.74 (68.06 – 74.19) | 7.57 (6.54 – 8.75) |
| Richer | 71.62 (68.92 – 74.19) | 7.17 (6.10 – 8.41) |
| Richest | 80.59 (78.39 – 82.62) | 9.62 (8.39 – 11.01) |
| **Children ever born** |  |  |
| No Child | 67.92 (65.75 – 70.00) | 4.05 (3.41 – 4.79) |
| 1-2 child | 74.40 (72.57 – 76.15) | 9.02 (8.24 – 9.87) |
| >2 Child | 69.17 (66.71 – 71.53) | 8.16 (7.00 – 9.49) |
| **Place of Residence** |  |  |
| Urban | 77.82 (75.23 – 80.21) | 8.99 (7.91 – 10.20) |
| Rural | 68.67 (66.46 – 70.81) | 6.62 (6.01 – 7.29) |
| **Place of regions** |  |  |
| Barisal | 69.43 (64.16 – 74.24) | 10.43 (8.45 – 12.82) |
| Chattogram | 67.42 (63.43 – 71.17) | 5.05 (4.11 – 6.19) |
| Dhaka | 76.80 (72.29 – 80.77) | 8.19 (6.80 – 9.84) |
| Khulna | 63.87 (58.70 – 68.73) | 7.01 (5.80 – 8.46) |
| Mymensingh | 77.86 (72.51 – 82.42) | 3.59 (2.74 – 4.69) |
| Rajshahi | 69.36 (64.63 – 73.71) | 7.59 (6.51 – 8.83) |
| Rangpur | 74.42 (70.44 – 78.03) | 10.64 (8.95 – 12.60) |
| Sylhet | 65.23 (58.89 – 71.07) | 5.76 (4.47 – 7.40) |
| **Community level illiteracy** |  |  |
| Low | 79.10 (74.73 – 82.88) | 10.03 (8.49 – 11.80) |
| Moderate | 70.72 (68.52 – 72.83) | 7.24 (6.57 – 7.97) |
| High | 67.94 (63.65 – 71.95) | 5.72 (4.66 – 6.99) |
| **Community level Poverty** |  |  |
| High poverty | 70.87 (68.28 – 73.33) | 6.33 (5.62 – 7.12) |
| Moderate poverty | 68.48 (64.46 – 72.24) | 6.47 (5.57 – 7.51) |
| Low poverty | 69.16 (64.83 – 73.18) | 9.61 (8.09 – 11.39) |
| Richest Community | 78.69 (74.78 – 82.14) | 9.54 (7.91 – 11.47) |

The factors associated with the MR service knowledge and use of MR were determined using the multilevel Poisson regression model. Five different models were run separately. The summary of each model is presented in Table 3. The model with the lowest intra-class correlation and AIC and BIC are the best model. Full model, for both the MR service knowledge and use of MR service, met this condition. We found around 16% difference of MR service knowledge across clusters which reduced to 9% once individual, household and community level factors were adjusted. Similarly, there were 22.10% difference of MR service use was found across cluster which declined to 13.10% following the adjustment of individual, household, and community level factors.

**Table 3:** Results from random intercept model (measure of variation) for menstrual regulation knowledge and menstrual regulation use at cluster level using multilevel Poisson regression analysis.

|  | Null model | Individual level model | Household level model | Community level model | Full model |
| --- | --- | --- | --- | --- | --- |
| **Menstrual regulation knowledge** |  |  |  |  |  |
| Intra-class correlation | 16.10% | 14.10% | 13.28% | 12.10% | 9.40% |
| Variance of the random intercept | 0.70 | 0.64 | 0.57 | 0.48 | 0.40 |
| AIC | 38186.7 | 37984.4 | 35100.4 | 38128.7 | 34973.7 |
| BIC | 38202.5 | 38056.01 | 35317.7 | 38246.7 | 35247.4 |
| **Menstrual regulation service use** |  |  |  |  |  |
| Intra-class correlation | 22.10% | 18.10% | 16.12% | 15.42% | 13.20% |
| Variance of the random intercept | 0.54 | 0.46 | 0.32 | 0.28 | 0.16 |
| AIC | 7551.04 | 7430.83 | 6964.96 | 7459.08 | 6854.04 |
| BIC | 7536.21 | 7499.09 | 7177.53 | 7572.86 | 7116.72 |

The results of the full models, for both the MR service knowledge and use of MR service, are presented in Table 4 while results of all models are presented in supplementary tables 1 and 2. The likelihoods of MR service knowledge were found to be higher among women aged 20-34 years (PR, 1.26, 95% CI, 1.20-1.33) and ≥35 years (PR, 1.30, 95% CI, 1.23-1.38) as compared to the women aged 15-19 years. Knowledge about MR service was found to be increased to around 6-16% among women with increased education from primary education. Alternatively, around 10% lower knowledge of MR service (PR, 0.90, 95% CI, 0.86-0.93) was found among illiterate women. Similar associations were reported for women’s partner education. We found around 4-9% lower knowledge of MR service among women with poorest and poorer socio-economic status as compared to the women with middle wealth quintile. Around 5-6% higher likelihoods of MR knowledge were found among women with increased number of children than among women who have no children. Rural women were 12% (PR, 0.88, 95% CI, 0.84-0.94) less likely to know about MR service as compared to the urban women. Women in the Mymensingh and Rangpur divisions were 10-17% more likely to know about MR service as compared to the women in the Barishal division. Knowledge about MR service was found to be increased with the increased level of illiteracy and decreased of community level poverty.

**Table 4:** Factors associated with knowledge about menstrual regulation service and menstrual regulation service use using the multilevel Poisson regression model

| Characteristics | Menstrual regulation knowledge, PR, (95% CI) | Menstrual regulation use, PR, (95% CI) |
| --- | --- | --- |
| **Age of respondents** |  |  |
| 15-19 years | 1.00 | 1.00 |
| 20-34 years | 1.26 (1.20 – 1.33)** | 3.13 (1.85 – 5.28)** |
| ≥35 years | 1.30 (1.23 – 1.38)** | 4.23 (2.44 – 7.32)** |
| **Respondent educational level** |  |  |
| No education | 0.90 (0.86 – 0.93)** | 0.85 (0.68 – 1.08) |
| Primary education | 1.00 | 1.00 |
| Secondary education | 1.06 (1.03 – 1.09)** | 0.85 (0.68 – 1.07) |
| Higher education | 1.16 (1.11 – 1.20)** | 0.86 (0.65 – 1.14) |
| **Age at first marriage** |  |  |
| Marriage (10-17 years) | 1.00 | 1.00 |
| 18 years and above | 0.98 (0.97 – 1.01) | 0.85 (0.73 – 1.01) |
| **Respondent currently working status** |  |  |
| No | 1.00 | 1.00 |
| Yes | 1.08 (1.05 – 1.11)** | 1.07 (0.73 – 1.00) |
| **Husband educational level** |  |  |
| No education | 0.96 (0.94 – 0.99)** | 0.90 (0.73 – 1.11) |
| Primary education | 1.00 | 1.00 |
| Secondary education | 1.04 (1.01 – 1.06)** | 1.18 (0.98 – 1.41) |
| Higher education | 1.06 (1.02 – 1.09)** | 1.05 (0.81 – 1.35) |
| **Husband occupational status** |  |  |
| Agriculture | 1.00 | 1.00 |
| Physical workers | 1.03 (1.00 – 1.05)* | 1.24 (1.03 – 1.50)* |
| Service | 1.04 (0.99 – 1.08) | 1.33 (0.98 – 1.82) |
| Business | 1.05 (1.00 – 1.08)** | 1.39 (1.13 – 1.70)** |
| Others | 1.00 (0.79 – 1.27) | 0.97 (0.23 – 4.01) |
| **Wealth Index** |  |  |
| Poorest | 0.91 (0.88 – 0.95)** | 0.71 (0.54 – 0.93)** |
| Poorer | 0.96 (0.93 – 0.99)* | 0.95 (0.75 – 1.21) |
| Middle | 1.00 | 1.00 |
| Richer | 0.99 (0.96 – 1.02) | 0.88 (0.70 – 1.29) |
| Richest | 1.04 (0.99 – 1.08) | 1.00 (0.78 – 1.29) |
| **Children ever born** |  |  |
| No Child | 1.00 | 1.00 |
| 1-2 child | 1.06 (1.03 – 1.09)** | 1.67 (1.37 – 2.04)** |
| >2 Child | 1.05 (1.00 – 1.08)** | 1.64 (1.27 – 2.14)** |
| **Place of residence** |  |  |
| Urban | 1.00 | 1.00 |
| Rural | 0.88 (0.84 – 0.94)** | 0.96 (0.79 – 1.16) |
| **Place of Regions** |  |  |
| Barisal | 1.00 | 1.00 |
| Chattogram | 0.98 (0.91 – 1.04) | 0.47 (0.35 – 0.62)** |
| Dhaka | 1.12 (1.02 – 1.23)* | 0.76 (0.57 – 1.01) |
| Khulna | 0.93 (0.84 – 1.02) | 0.68 (0.51 – 0.90)** |
| Mymensingh | 1.17 (1.07 – 1.27)** | 0.42 (0.30 – 0.58)** |
| Rajshahi | 1.04 (0.94 – 1.13) | 0.84 (0.65 – 1.07) |
| Rangpur | 1.10 (1.01 – 1.19)** | 1.25 (0.98 – 1.60) |
| Sylhet | 1.01 (0.91 – 1.13) | 0.63 (0.45 – 0.88)** |
| **Community level Poverty** |  |  |
| High poverty | 1.00 | 1.00 |
| Moderate poverty | 0.91 (0.86 – 0.97)** | 0.99 (0.81 – 1.22)** |
| Low poverty | 0.87 (0.81 – 0.94)** | 1.41 (1.07 – 1.85)** |
| Middle and richest Community | 0.86 (0.78 – 0.96)* | 1.28 (0.92 – 1.79) |
| **Community level illiteracy** |  |  |
| Low | 1.00 | 1.00 |
| Moderate | 0.92 (0.87 – 0.98)** | 0.82 (0.68 – 0.99)* |
| High | 0.89 (0.82 – 0.97)** | 0.67 (0.51 – 0.88)** |

The MR service use was found to be increased to 3.13 times (95% CI, 1.85-5.28) and 4.23 times (95% CI, 2.44-7.32) among women aged 20-34 years and ≥35 years, respectively, as compared to the women aged 15-19 years. Likelihoods of MR service use was also found higher among women whose husbands were occupations were either physical worker (PR, 1.24, 95% CI, 1.03-1.50) or business (PR, 1.39, 95% CI, 1.13-1.70) as compared to the women whose husbands were engaged with agricultural work. As compared to the women with middle wealth quintile, women with poorest wealth quintile were 29% (PR, 0.71, 95% CI, 0.54-0.93) less likely to use MR service. Around 64-67% higher likelihoods of MR service use were found among women with increased number of children than women with no children. Women resided in the Khulna (PR, 0.68, 95% CI, 0.51-0.90) and Mymensingh (PR, 0.42, 95% CI, 0.30-0.58) divisions were less likely to use MR as compared to the women resided in the Barishal division. Use of MR service was also found to be increased with the decreased of community level poverty and increase of community level illiteracy.

## Discussion

In this study, we explored the knowledge and prevalence of MR service use in Bangladesh using most recent 2017 BDHS data. We found around 71.28% of the total women analysed know about this service while 7.36% of the women used this service within three years prior to the survey. Knowledge about MR service was found higher among higher aged women, higher educated women, women who engaged in income generating employment, women whose husbands were either work as physical worker or do business, women with lower socio-economic quintile and women with increased number of children. Rural women and women who resided in the community with low poverty and higher illiteracy were less likely to know about MR service. MR service use was found higher among higher aged women, women whose husbands were either physical workers or businessmen, women who having increased number of children and resided in the community with lower poverty. Lower use of MR service was found among women resided in the Chattogram, Khulna and Mymensingh divisions and women resided in the community with increased illiteracy. Findings will help the policy makers to make evidence-based policy and program to increase knowledge of MR service and its use whenever necessary.

We found higher knowledge and use of MR among increased aged women which are consistent with the findings of previous studies conducted in other LMICs (Alam & Sultan, 2019; Park et al., 2020; Reiss et al., 2019; Singh et al., 2017b). Lower aged women are more likely to be influenced by social stigma which might be linked with such lower use of MR. Moreover, increased aged women were less likely to use contraception or depend on less effective or traditional contraception’s (Blake et al., 2007; Zahan & Feng, 2020). Also, they have already had desired number of children, so that any pregnancy is treated as unintended. Therefore, they consider MR service use (Bose S & Trent K 2006; Mote et al., 2010). Alternatively, if any unintended pregnancy occurred among lower aged women, who do not have required number of children, they may compromise with the time and continue the pregnancy to live births.

As like other studies in LMICs, in this study we found higher knowledge and use of MR among women with increased education (Alam & Sultan, 2019; Ankara, 2017; DaVanzo et al., 2013; Mote et al., 2010; Rana et al., 2019). Educated women were more likely to know about the reproductive and family planning services options either as part of their academic education, higher community level engagement, and higher exposure of mass medica. Also, they have better decision-making autonomy over their reproductive life and know more about the importance of lower number of children (DaVanzo et al., 2013). This association is also justified by the higher use of MR service among with increased number of children as found in this study which is also similar to the previous studies in LMICs (Alam & Sultan, 2019; Aleni et al., 2020; Allotey et al., 2021; Dickson et al., 2018). Together these lead to higher use of MR service among women with increased education.

We also reported knowledge and use of MR services were different across divisions of Bangladesh and urban vs rural which are consistent with the available studies of LMICs (Chae et al., 2017; Kant et al., 2015; Kapil Ahmed et al., 2005; Rana et al., 2019; Yassin K M 2000). These variations could be explained by community level social stigma, and community level religious and cultural norms. Beside these, proximity of healthcare facility is also important as found in a recent study of Bangladesh (Bearak et al., 2020; Rana et al., 2019).

Traditionally in Bangladesh, a community where people resided in is format with a combination of group of people with similar socio-economic status. Therefore, a majority or all of the people in lower poverty or illiterate community are either rich or educated. This direct higher knowledge and use of MR service among lower poverty and illiterate community as found in this study. In addition to these community level norms and social stigma, which are found more influential in the community with higher poverty and illiteracy, are also important for such associations (Ahmed & Ray, 2014; Ankara, 2017; Arambepola et al., 2016; Chae et al., 2017; Fusco et al., 2012; Johnston et al., 2010; SM Zahedul Islam, 2020; Vlassoff et al., 2012).

This study has several strengths and few limitations. In this study, we used most recent nationally representative data with a very higher sample size representative to all administrative divisions and urban and rural areas. We considered a range of individual-, household-, and community level factors and determined their linkages by using advanced statistical methods. Clusters level variations and sample weight were also considered in all analyses. Moreover, for the first time we calculated age-standardized prevalence of MR service knowledge and use. Therefore, the findings are mostly applicable for evidence-based decision and program making. However, the data analysed in this study were cross-sectional, therefore, the findings reported in this study were correlational only rather than casual. Moreover, since MR service use data were collected by asking directly to respondents without any validation, therefore, recall bias could be a major issue. Due to social and cultural norms, women may not give the accurate response on MR use status. However, any of such bias is likely to be random. In addition, health facility level factors, including MR service availability are important predictors of MR service knowledge and use of MR. However, we could not adjust this variable in our analysis due to lack of data. Another important limitation of this study the BDHS authority did not mention in which places MR services use taken of the women.

## Conclusion

Around 72% of the analysed women knew about MR while only 7% of them used MR within three years of the survey date. Several individual, household, and community level factors, including women’ age, education, number of children, women’s husbands’ age, occupation, place of residence, division and community status in response to poverty and illiteracy were found to be associated with MR service knowledge and use of MR in Bangladesh. Findings suggest requirement of policy and program in the community level to increase knowledge of MR and its use as an effective family planning services to reduce burden of unintended conception in Bangladesh. This should include awareness building program at the community level to encourage people to access MR service when the pregnancy was undesired.

## Data Availability

The datasets used and analyzed in this study are available from the Measure DHS website: https://dhsprogram.com/data/available-datasets.cfm?

## Acknowledgments

The authors thank MEASURE DHS for granting access to the BDHS 2017 data. The data is available on the website of the global DHS program, which could be accessed after getting registered with the MEASURES DHS. We wish to thank the director Aree Jampaklay, IPSR, MU, Thailand. The authors also thank to TWAS-IsDB and the editor and reviewers for their critical comments for the improvement of the manuscript.

## Authors’ contributions

MRA developed the study concepts. MRA and MNK analyzed the data. MRA drafted the manuscript. MRA, MNK and YS critically reviewed the manuscript. All authors have read and approved the final version of the paper.

## Data availability

The datasets used and analyzed in this study are available from the Measure DHS website: https://dhsprogram.com/data/available-datasets.cfm

